# Molecular signature of pediatric B-ALL determines outcomes post CD19 CAR-T cell therapy

**DOI:** 10.64898/2026.04.11.26350681

**Authors:** Aleksandra Oszer, Agata Pastorczak, Zuzanna Urbańska, Karolina Miarka, Paweł Marschollek, Monika Richert-Przygońska, Monika Mielcarek-Siedziuk, Christina Baggott, Liora Schultz, Jennifer Moon, Catherine Aftandilian, Jan Styczyński, Krzysztof Kałwak, Wojciech Młynarski, Kara L. Davis

## Abstract

Chimeric antigen receptor T-cell (CAR-T) therapy targeting CD19 has transformed outcomes for children with relapsed or refractory (R/R) B-cell acute lymphoblastic leukemia (B-ALL), yet the influence of molecular subtype on outcomes remains unclear. We evaluated the impact of cytogenetic and molecular signatures on complete response (CR), overall survival (OS), and leukemia-free survival (LFS) after CD19 CAR-T therapy in eighty-six pediatric patients with R/R B-ALL treated with tisagenlecleucel. CR was assessed 30 days after infusion. Cytogenetic data were available for 84 patients and molecular profiling for 62.

Survival analyses included 72 patients who received CD19 CAR-T as the sole cellular therapy. Seventy-seven patients achieved CR (89.5%). Pre-infusion bone marrow blasts of ≥20% were associated with lower CR rates (53.8% vs 95.9%, p<0.0001) and significantly reduced OS and LFS (both p<0.0001). Among molecular markers, RAS mutations correlated with inferior OS (p=0.0222) and LFS (0.0402). In multivariate analysis, bone marrow blasts >20% and RAS mutations independently predicted inferior OS. Post CAR-T, CD19 negative relapses showed almost twice higher prevalence of RAS mutations (66% vs 37.5%). These findings highlight RAS mutations as a key molecular predictor of outcome after CD19 CAR-T therapy and suggest emergence of unique risk stratification for patients receiving CD19-targeting therapy.

**Key Points:**

- **RAS mutations independently predict unfavorable survival after CAR-T CD19 in pediatric B-ALL**.
- **RAS mutations increase risk of CD19 negative relapse after CAR-T CD19 therapy in pediatric B-ALL**.

## Introduction

Chimeric antigen receptor T-cell (CAR-T) therapy directed against CD19 has transformed the treatment landscape for children with relapsed or refractory (R/R) B-cell acute lymphoblastic leukemia (B-ALL). Tisagenlecleucel (Kymriah), an autologous CD19-targeted CAR-T product, is the only therapy approved for pediatric B-ALL by both the U.S. Food and Drug Administration (FDA) and the European Medicines Agency (EMA).^1,2^ Clinical trials demonstrated high initial complete response (CR) rates of approximately 90% at day 30 after infusion,^3^ and the ELIANA study, with the longest available follow-up, reported 3-year event-free survival of 44% and overall survival (OS) of 63%.^4^ Despite these advances, a substantial proportion of patients relapse or require additional disease directed therapy.^5^

Although certain clinical parameters have been proposed to influence CAR-T efficacy,^3,6,7^ the contribution of underlying cytogenetic and molecular features remains insufficiently characterized, particularly in pediatric patients uniformly treated with a single licensed CAR-T product. Most prior studies combine heterogeneous CAR constructs, mixed age groups, or diverse disease subtypes, limiting the ability to evaluate biologic determinants of response.

To address this knowledge gap, we investigated the impact of cytogenetic and molecular signatures on response and survival in pediatric R/R B-ALL patients treated exclusively with tisagenlecleucel.

## Methods

Eighty-six pediatric patients (aged 1-25 years) with R/R B-ALL who received tisagenlecleucel were included. Patients were treated at two centers: Poland (n=42; March 2020-August 2024) and the United States (n=44; April 2017-September 2024). All patients or guardians provided written informed consent, and the study was approved by the Human Research Ethics Committee of the Medical University of Lodz and Medical University of Silesia and Stanford University (PCN/0022/KB1/90/XU/2021, KNW/0022/KB1/153/I/16/17, CCT5019). Clinical data were obtained retrospectively.

CR at day 30 post-infusion was assessed using PCR-based minimal residual disease (MRD) or next-generation sequencing (NGS) MRD,^8^ additionally FC-MRD was performed.^9^ Cytokine release syndrome (CRS) and neurotoxicity were graded using the Penn^10^ and ASTCT scales,^11,12^ respectively. Cytogenetic data were available for 84 patients. Molecular characterization, performed using RNA-sequencing, microarray profiling, and targeted next-generation sequencing panels was available for 62 patients (Supplemental Figure 1), detailed protocols are in Supplemental Methods.

OS and LFS analyses were restricted to 72 patients treated exclusively with CD19-directed tisagenlecleucel (Supplemental Figure 2).

CR rates were compared using Chi-square testing. OS and LFS were estimated with the Kaplan-Meier method and compared using the Gehan-Breslow-Wilcoxon test. Logistic regression evaluated predictors of CR, and multivariate Cox proportional hazards models assessed factors associated with OS and LFS. Analyses were conducted using GraphPad Prism 10.6.1, and R 4.5.1. Two-sided p<0.05 was considered statistically significant.

## Results and discussion

### Cohort characteristics and early response

Baseline clinical and biological characteristics of the 86 pediatric R/R B-ALL patients treated with tisagenlecleucel are summarized in Figure 1. Individual clinical courses are illustrated in a swimmer plot (Figure 1A) with distribution of recurrent genomic alterations among patients with available profiling shown in Figure 1B. Across the full cohort, 77 of 86 patients (89.5%) achieved CR at day 30. CR rates were comparable across sex, age, B-ALL immunophenotypic subtype, number of prior relapses, prior blinatumomab or inotuzumab ozogamicin exposure, prior hematopoietic stem cell transplantation (HSCT), neurotoxicity grade, cytogenetic risk categories, and molecular subgroups. Consistent with clinical experience and prior reports^13^, higher morphologic marrow blast burden at infusion strongly associated with nonresponse: patients with bone marrow (BM) blasts ≥20% had a substantially lower CR rate than those with <20% (53.8% vs 95.9%, p<0.0001). Notably, trends toward lower CR rates were observed in subgroups with *PAX5* rearrangements (*PAX5r*; 60% CR, p=0.0866) and *KMT2A* rearrangements (*KMT2Ar*; 50% CR, p=0.0551), but these did not reach statistical significance (Figure 2; Supplemental Table 1), likely reflecting limited sample size.

**Figure 1.**
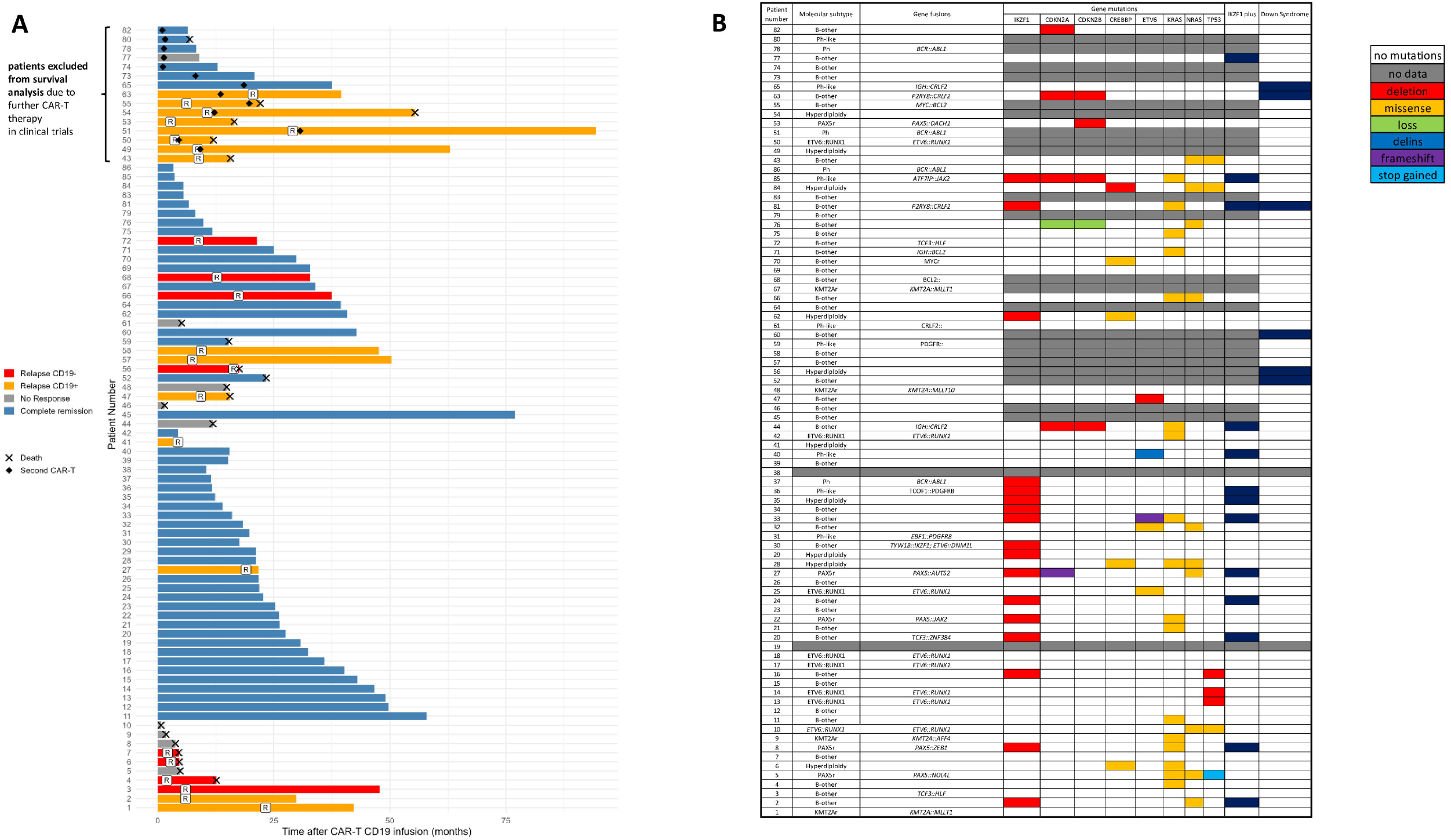
Characteristics of pediatric patients treated with CAR-T CD19 therapy. (A)Swimmer plot depicting individual patient timelines. Background colors indicate response status: non-responders, sustained remission, relapse or death. (B)Gene mutation profiles per patient. Each color represents a specific mutation type; gray indicates unavailable data. IKZF1 Plus is defined as the presence of an IKZF1 deletion in acute lymphoblastic leukemia accompanied by concomitant deletions in CDKN2A/B, PAX5, or PAR1, in the absence of ERG deletion, and is associated with adverse prognosis.^22^

**Figure 2.**
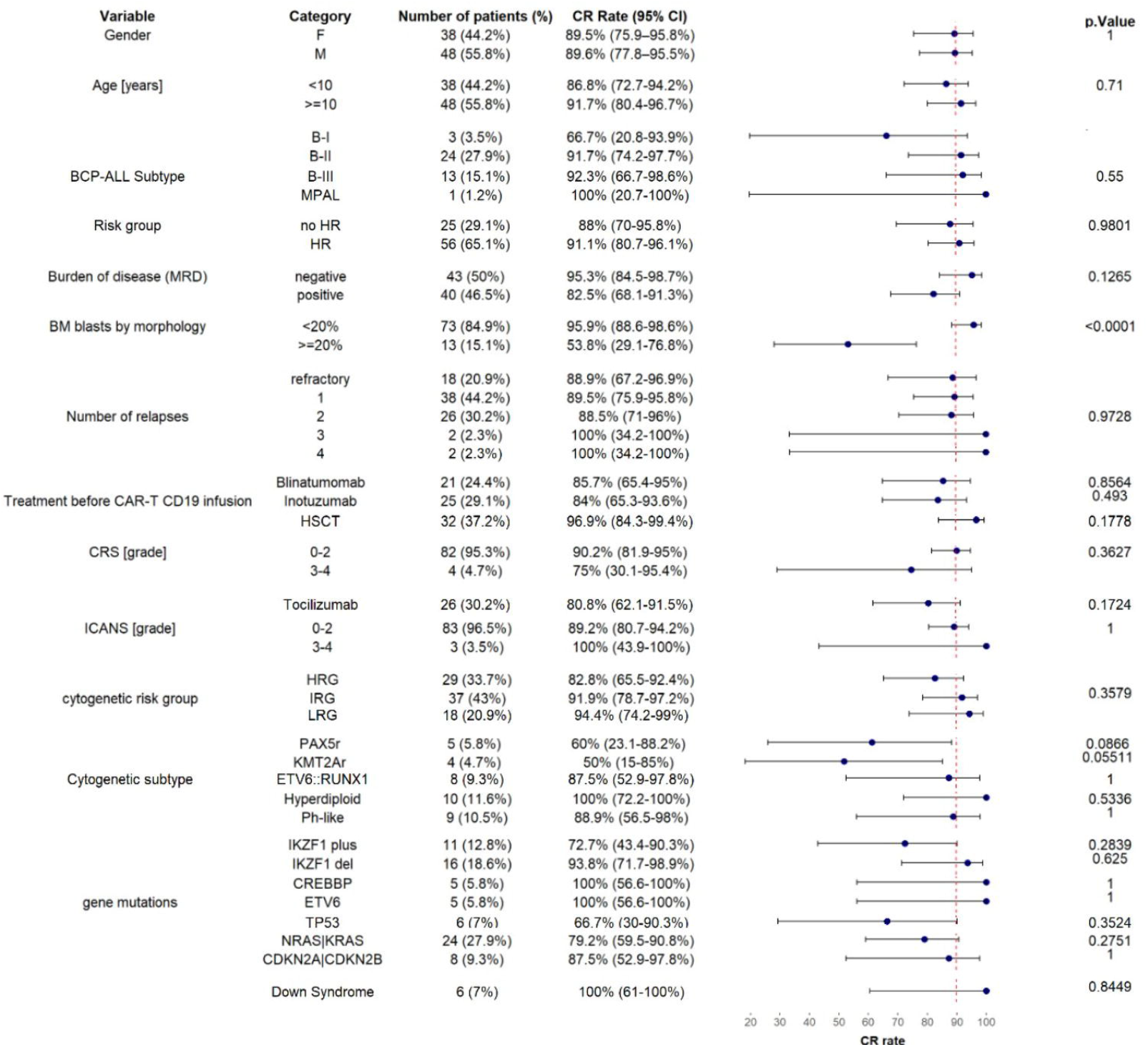
Univariate analysis of CR rates (30 days post-CAR-T CD19 infusion) across different subgroups. Among the 86 patients who received CAR-T CD19 cell therapy (tisagenlecleucel), those with bone marrow (BM) blasts >20% had significantly lower CR rates compared to those with BM blasts ≤20% (53.8% vs. 95.9%, p<0.0001). CR rates according to clinical and biological features were analyzed using the Chi-square test, and results were visualized using forest plots.

### Longer-term outcomes after tisagenlecleucel

Overall and leukemia-free survival were analyzed in the subset treated exclusively with CD19-directed tisagenlecleucel, excluding patients who received additional CAR products (Supplemental Figure 2). Two-year OS and LFS were 74.4% and 62.2%, respectively (Supplemental Figure 3A-B). Baseline BM blasts ≥20% was associated with markedly inferior outcomes (p<0.0001 for both OS and LFS; Figure 3A-B; Supplemental Table 2), reinforcing the clinical importance of disease burden at infusion.^14,15^

**Figure 3.**
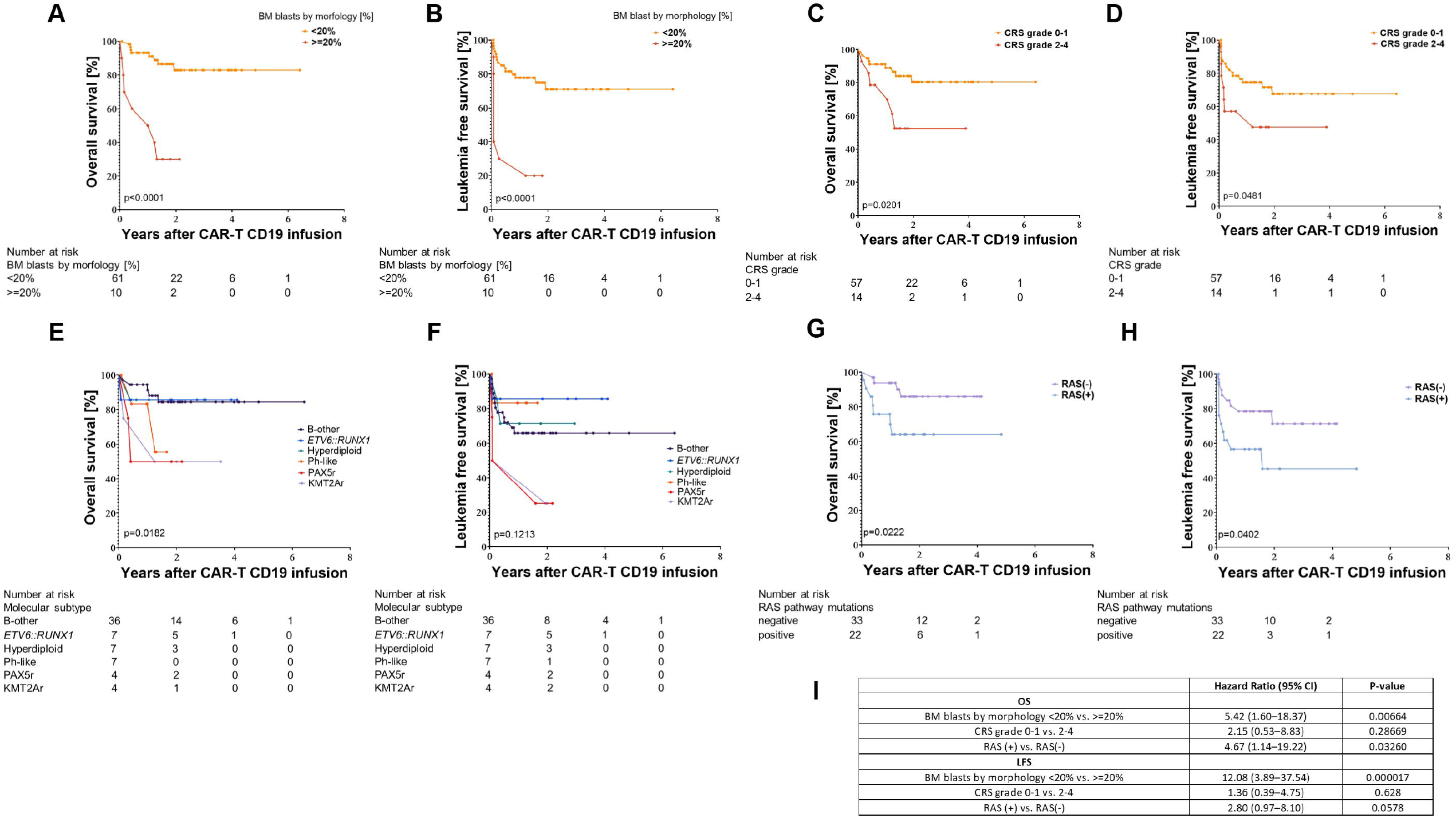
Impact of clinical and molecular factors on OS and LFS among 71 patients receiving CAR-T CD19 therapy. (A-B) OS and LFS stratified by BM blasts >20% vs. ≤20%. (C-D) Outcomes stratified by cytokine release syndrome (CRS) grade ≥2 vs. <2. (E-F) Outcomes by cytogenetic subgroup. (G-H) Outcomes based on RAS mutation. (I) Multivariate analysis on OS and LFS by subgroup in the patients who received CAR-T CD19 therapy, using multivariate Cox proportional hazards regression models.

CRS severity also correlated with outcome: patients with CRS grade ≥2 had reduced OS (p=0.0201) and LFS (p=0.0481) compared with CRS grade 0-1 (Figure 3C-D). Because CRS severity increases with leukemia burden, these findings likely reflect combined effects of disease biology and inflammatory toxicity.^16^ Higher blast burden correlated with CRS grade 2-4 (p=0.0006), and tocilizumab use (p=0.0090; Supplemental Table 3). Neurotoxicity grade was not significantly associated with survival, although the number of high-grade ICANS cases was small.

### Cytogenetic signatures associated with outcome

Cytogenetic risk groups (high, intermediate, low) did not differ significantly in OS or LFS (Supplemental Table 2), aligning with recent larger datasets reporting comparable CAR-T outcomes across traditional cytogenetic strata.^7^

In contrast, specific genetic lesions showed potential clinical significance. *PAX5r*(+) and *KMT2Ar* (+) patients had significantly reduced OS (p=0.0182) and a trend toward reduced LFS (p=0.1213) (Figure 3E-F).

### RAS signature as independent predictor

RAS pathway alterations, both NRAS and/or KRAS was associated with inferior OS (p=0.0222) and LFS (p=0.0402) (Figure 3G-H).

In multivariable Cox models, BM blasts ≥20% and RAS remained the only independent predictors of inferior survival outcomes. BM blasts ≥20% strongly predicted inferior OS (HR 5.42 ; p=0.0066) and LFS (HR 12.08; p<0.001). RAS mutations independently predicted OS (HR 4.67; p=0.0326) and a trend toward reduced LFS (HR 2.8; p=0.0578). CRS status did not retain statistical significance in adjusted models (OS HR 2.15, p=0.2866; LFS HR 1.36, p=0.628, Figure 3I).

Additionally, 66% (6/9) of CD19 negative relapse after CAR-T CD19 had RAS mutations, while 37.5% (3/8) of CD19-positive relapse had RAS mutation.

Given prior reports that clonal RAS mutations predict unfavorable outcomes in R/R B-ALL^17^, we compared the impact of clonal vs. subclonal RAS (<25% variant allele frequency (VAF)) but observed no significant differences in OS or LFS (Supplement Figure 4).

Taken together, these findings identify RAS mutations as a robust molecular marker of poor outcome following tisagenlecleucel in pediatric B-ALL, independent of leukemia burden and other known clinical variables.

To evaluate potential bias from incomplete molecular testing and additiongal risk factors in RAS(+), we compared baseline clinical features of patients with and without available molecular data, and RAS(+) vs RAS(-). No significant differences were observed across leukemia burden, toxicity, pretreatment exposures, or timing from diagnosis/relapse to infusion (Supplemental Table 4, 5), indicating that missing molecular data or RAS mutation were not systematically enriched for any risk group.

### Comparison with existing literature

Our findings align with and further extend the observations of Leahy et al.,^7^ who reported similar CR, OS, and RFS rates across cytogenetic risk strata in a large cohort of pediatric/young adult patients receiving tisagenlecleucel, CTL019, or huCART19. In their analysis, traditional high-risk lesions, including *KMT2A* rearrangements, Ph+, Ph-like, and hypodiploidy did not independently predict post-CAR-T relapse. Our study is consistent with this broader literature by showing no significant prognostic impact of cytogenetic risk groups. However, unlike prior reports, our dataset identifies *PAX5* rearrangements, that have been linked to reduced CD58 expression and diminished blinatumomab response,^18^ underscoring its potential relevance. Considering the low percentage representation of *PAX5r* cases and the diverse profile of *PAX5* gene partners, this relationship requires further in-depth analysis, especially, since RAS lesions co-occurred in 4 out of 5 patients with *PAX5r* in our study group.

Previous studies highlighted TP53 mutation and high leukemia burden as dominant drivers of failure after CAR-T CD19^3,6^. In our cohort, TP53 (n=6, 7% of cohort) did not significantly influence CR, OS, or LFS. In addition, Pan et al.^6^ report the incidence of RAS mutations in that patients cohort but without providing further analysis of their prognostic significance. Our data reveal the association of RAS mutations with the incidence of post-CAR CD19-negative relapse. RAS mutations may designate patients eligible for MEK inhibitor^19,20^ after CAR-T CD19 therapy preventing CD19 negative relapse.

### Interpretation and implications

This study supports three clinically significant conclusions.

First, tisagenlecleucel achieves high early remission rates (89.5%) in pediatric R/R B-ALL, with most baseline clinical variables exerting little influence on day-30 response.

Second, leukemia burden remains a powerful determinant of both remission and survival, reinforcing the clinical priority of achieving low disease burden prior to CAR-T infusion.

Third, RAS mutations represent high-risk molecular marker associated with inferior outcomes that persist after adjustment for disease burden. Given the significant role of RAS signaling in B-cell lineage development specification,^21^ RAS-altered leukemias may possess intrinsic features that limit CAR-T cytotoxicity or facilitate early CD19-negative relapse.

### Study limitations and future directions

This study is limited by its retrospective design, incomplete molecular profiling in some patients, and modest sample sizes for less frequent lesions. However, the use of a single, FDA/EMA-approved CAR-T product and a pediatric-only cohort strengthens interpretability and reduces confounding from product or age heterogeneity.

## Conclusion

Overall, these data highlight RAS mutations as a clinically meaningful molecular predictor of outcome following CD19 CAR-T therapy in pediatric B-ALL, including an increased risk of CD19 negative relapse. Incorporating RAS-pathway alterations into post-CAR-T risk stratification may enable more personalized consolidation strategies and improved long-term survival.

## Data Availability

Original data and protocols are available on request from the corresponding author, Aleksandra Oszer.

## Acknowledgments

We express our sincere gratitude to the children and families whose clinical histories made this retrospective study possible. We deeply appreciate the dedicated healthcare professionals involved in their care. In particular, we thank the clinical teams from the Division of Hematology, Oncology, Stem Cell Transplant, and Regenerative Medicine, Department of Pediatrics, Stanford University (Stanford, CA, USA); the Department of Pediatric Bone Marrow Transplantation, Oncology, and Hematology at Wroclaw Medical University (Wroclaw, Poland); and the Department of Pediatric Hematology, Oncology, Immunology and Transplantology, Nicolaus Copernicus University Torun, Collegium Medicum (Bydgoszcz, Poland) for their expert clinical management, diligent documentation, and longstanding commitment to the patients.

We are also grateful to the medical, laboratory, and administrative staff whose assistance in record retrieval, data verification, and clinical support contributed significantly to the completion of this study.

This project was financed by the National Science Centre, Poland (PRELUDIUM 2023/49/N/NZ6/02818), and NAWA - Polish National Agency for Academic Exchange in cooperation with Medical Research Agency under the Walczak Programme (BPN/WAL/2023/1/00008). Genetic studies were partially supported by the cALL-POL project financed by the Medical Research Agency.

## Authorship Contributions

A.O., A.P., K.L.D., and W.M. contributed to the conception, design, and overall planning of the study. A.O. performed data curation, formal analysis, secured funding, and wrote the initial draft. A.P. contributed substantially to methodology, data interpretation, and writing-review & editing. Z.U. and K.M. conducted laboratory analyses and contributed to data processing and verification. P.M., M.R.-P., M.M.-S., C.B., L.S., J.M., C.A., J.S., and K.K. contributed to clinical data acquisition and clinical expertise. W.M. and K.L.D. provided supervision, critical manuscript revision, and additional funding acquisition.

All authors read and approved the final manuscript.

