## Supplemental data for "Molecular signature of pediatric B-ALL determines outcomes post CD19 CAR-T cell therapy"

### Supplemental Methods

#### A Single nucleotide polymorphism (SNP) arrays

Copy-number variation analysis was performed using CytoScan HD array (2 670 000 markers, including 750 000 single-nucleotide polymorphism (SNP) and 1 900 000 nonpolymorphic copy number variant (CNV) markers); (Applied Biosystems, Thermo Fisher, Waltham, MA, USA). All laboratory procedures were conducted according to the manufacturer's protocols and standardized based on the AIEOP-BFM array screening strategy recommendations for microdeletion and microduplication assessments. In total, 250 ng of the genomic DNA was processed according to the manufacturers' protocol following the steps of digestion by NspI, amplification using a ligation-mediated PCR with adapters linked to the restriction fragments, purification of PCR products using magnetic beads, fragmentation using DNase I, labeling with Terminal deoxynucleotidyl transferase (TdT) and hybridization overnight (16–18 h) to a 49-format array. After incubation samples were washed and stained with the use of a GeneChip Fluidics Station 450 and were scanned by GeneChip Scanner 3000. To generate the CEL files that include the intensity of probe signals the GeneChip Command Console Software (Thermo Fisher Scientific) was used. Then the Chromosome Analysis Suite v 4.5 software (ChAS, Thermo Fisher Scientific) converted CEL to CYCHP files containing information on copy number, loss of heterozygosity (LOH), mosaicism, and genotype calls. Copy number state and SNP genotypes were called using the Hidden Markov Model (HMM) algorithm and the Bayesian Robust Linear Model with the Mahalanobis distance classifier (BRLMM) algorithm, respectively. The intensity ratio of each SNP and CN probe in the DNA test provides a relative copy number (log2 ratio: log2sample − log2reference) which is normalized concerning a reference. The Reference Model File contains 380 samples, 284 from HapMap and 96 from BioServe Biotechnologies (BioServe Biotechnologies, Beltsville, MD, USA). Determination of log2 ratio indicated if there is a gain or loss of genetic material. The threshold levels of log2 ratio ≥0.5 and ≤0.5 were used for the categorization of the altered chromosomal regions as CNV gains and losses, respectively.

**B RNA sequencing array**

RNA sequencing was performed using the TruSight RNA Pan-Cancer panel (Illumina, San Diego, CA), which contains 1,385 cancer genes and enables fusion calling and variant detection within the panel. Fifty nanograms of RNA were processed according to the manufacturer's protocol and sequenced on a NextSeq 550 system (Illumina, San Diego, CA) using the NextSeq Reagent Kit v2.5 (300 cycles) with a PE NextSeq Flow Cell. Data analysis was performed using the Illumina BaseSpace apps: DRAGEN RNA (version 4.0.4), fusion calling by TopHat-Fusion2, and RNA-seq Alignment (version 2.0.2, read mapping on the hg19 reference genome by STAR3, fusion calling by Manta4), using standard settings (https://basespace.illumina.com/apps). Fusion transcripts with a low number of split reads (<10) were excluded as likely false positives. Raw data of sequence variants were converted to variant call format (vcf) files and analysed in Variant Studio software v.4.0.

The list of genes included in targeted RNA Sequencing:

| | | | | | | |
|---|---|---|---|---|---|---|
| *ABCC3* | *ABI1* | *ABL1* | *ABL2* | *ABLIM1* | *ACACA* | *ACE* |
| *ACER1* | *ACKR3* | *ACSBG1* | *ACSL3* | *ACSL6* | *ACVR1B* | *ACVR1C* |
| *ACVR2A* | *ADD3* | *ADM* | *AFF1* | *AFF3* | *AFF4* | *AGR3* |
| *AHCYL1* | *AHI1* | *AHR* | *AHRR* | *AIP* | *AK2* | *AK5* |
| *AKAP12* | *AKAP6* | *AKAP9* | *AKR1C3* | *AKT1* | *AKT2* | *AKT3* |
| *ALDH1A1* | *ALDH2* | *ALDOC* | *ALK* | *AMER1* | *AMH* | *ANGPT1* |
| *ANKRD28* | *ANLN* | *APC* | *APH1A* | *APLP2* | *APOD* | *AR* |
| *ARAF* | *ARFRP1* | *ARHGAP20* | *ARHGAP26* | *ARHGEF12* | *ARHGEF7* | *ARID1A* |
| *ARID2* | *ARIH2* | *ARNT* | *ARRDC4* | *ASMTL* | *ASPH* | *ASPSCR1* |
| *ASTN2* | *ASXL1* | *ATF1* | *ATF3* | *ATG13* | *ATG5* | *ATIC* |
| *ATL1* | *ATM* | *ATP1B4* | *ATP8A2* | *ATR* | *ATRNL1* | *ATRX* |

| | | | | | | |
|---|---|---|---|---|---|---|
| *AURKA* | *AURKB* | *AUTS2* | *AXIN1* | *AXL* | *BACH1* | *BACH2* |
| *BAG4* | *BAIAP2L1* | *BAP1* | *BARD1* | *BAX* | *BAZ2A* | *BCAS3* |
| *BCAS4* | *BCL10* | *BCL11A* | *BCL11B* | *BCL2* | *BCL2A1* | *BCL2L1* |
| *BCL2L2* | *BCL3* | *BCL6* | *BCL7A* | *BCL9* | *BCOR* | *BCORL1* |
| *BCR* | *BDNF* | *BHLHE22* | *BICC1* | *BIN1* | *BIRC3* | *BIRC6* |
| *BLM* | *BMP4* | *BMPR1A* | *BRAF* | *BRCA1* | *BRCA2* | *BRD1* |
| *BRD3* | *BRD4* | *BRIP1* | *BRSK1* | *BRWD3* | *BTBD18* | *BTG1* |
| *BTG2* | *BTK* | *BTLA* | *BUB1B* | *C11orf1* | *C11orf30* | *C11orf54* |
| *C11orf95* | *C2CD2L* | *C2orf44* | *C3orf27* | *CACNA1F* | *CACNA1G* | *CACNA2D3* |
| *CAD* | *CALR* | *CAMK2A* | *CAMK2B* | *CAMK2G* | *CAMTA1* | *CANT1* |
| *CAPRIN1* | *CAPZB* | *CARD11* | *CARM1* | *CARS* | *CASC5* | *CASP3* |
| *CASP7* | *CASP8* | *CAV1* | *CBFA2T3* | *CBFB* | *CBL* | *CBLB* |
| *CBLC* | *CCAR2* | *CCDC28A* | *CCDC6* | *CCDC88C* | *CCK* | *CCL2* |
| *CCNA2* | *CCNB1IP1* | *CCNB3* | *CCND1* | *CCND2* | *CCND3* | *CCNE1* |
| *CCNG1* | *CCT6B* | *CD19* | *CD22* | *CD274* | *CD28* | *CD36* |
| *CD44* | *CD58* | *CD70* | *CD74* | *CD79A* | *CD79B* | *CD8A* |
| *CDC14A* | *CDC14B* | *CDC25A* | *CDC25C* | *CDC42* | *CDC73* | *CDH1* |

| | | | | | | |
|---|---|---|---|---|---|---|
| *CDH11* | *CDK1* | *CDK12* | *CDK2* | *CDK4* | *CDK5RAP2* | *CDK6* |
| *CDK7* | *CDK8* | *CDK9* | *CDKL5* | *CDKN1A* | *CDKN1B* | *CDKN1C* |
| *CDKN2A* | *CDKN2B* | *CDKN2C* | *CDKN2D* | *CDX1* | *CDX2* | *CEBPA* |
| *CEBPB* | *CEBPD* | *CEBPE* | *CENPF* | *CENPU* | *CEP170B* | *CEP57* |
| *CEP85L* | *CHCHD7* | *CHD2* | *CHD6* | *CHEK1* | *CHEK2* | *CHIC2* |
| *CHL1* | *CHMP2B* | *CHN1* | *CHST11* | *CHUK* | *CIC* | *CIITA* |
| *CIRH1A* | *CIT* | *CKB* | *CKS1B* | *CLP1* | *CLTA* | *CLTC* |
| *CLTCL1* | *CMKLR1* | *CNBP* | *CNOT2* | *CNTN1* | *CNTRL* | *COG5* |
| *COL11A1* | *COL1A1* | *COL1A2* | *COL3A1* | *COL6A3* | *COL9A3* | *COMMD1* |
| *COX6C* | *CPNE1* | *CPS1* | *CPSF6* | *CRADD* | *CREB1* | *CREB3L1* |
| *CREB3L2* | *CREBBP* | *CRKL* | *CRLF2* | *CRTC1* | *CRTC3* | *CSF1* |
| *CSF1R* | *CSF3* | *CSF3R* | *CSNK1G2* | *CSNK2A1* | *CTCF* | *CTDSP2* |
| *CTLA4* | *CTNNA1* | *CTNNB1* | *CTNND2* | *CTRB1* | *CTSA* | *CUX1* |
| *CXCL8* | *CXCR4* | *CXXC4* | *CYFIP2* | *CYLD* | *CYP1B1* | *CYP2C19* |
| *DAB2IP* | *DACH1* | *DACH2* | *DAXX* | *DCLK2* | *DCN* | *DDB2* |
| *DDIT3* | *DDR2* | *DDX10* | *DDX20* | *DDX39B* | *DDX3X* | *DDX5* |
| *DDX6* | *DEK* | *DGKB* | *DGKI* | *DGKZ* | *DICER1* | *DIRAS3* |

| | | | | | | |
|---|---|---|---|---|---|---|
| *DIS3L2* | *DKK1* | *DKK2* | *DKK4* | *DLEC1* | *DLL1* | *DLL3* |
| *DLL4* | *DMRT1* | *DMRTA2* | *DNAJB1* | *DNM1* | *DNM2* | *DNM3* |
| *DNMT1* | *DNMT3A* | *DOCK1* | *DOT1L* | *DPM1* | *DPYD* | *DST* |
| *DTX1* | *DTX4* | *DUSP2* | *DUSP22* | *DUSP26* | *DUSP9* | *DUX4* |
| *E2F1* | *EBF1* | *ECT2L* | *EDIL3* | *EDNRB* | *EED* | *EEFSEC* |
| *EGF* | *EGFR* | *EGR1* | *EGR2* | *EGR3* | *EGR4* | *EIF4A2* |
| *EIF4E* | *ELF4* | *ELK4* | *ELL* | *ELN* | *ELOVL2* | *ELP2* |
| *EML1* | *EML4* | *ENPP2* | *EP300* | *EP400* | *EPC1* | *EPCAM* |
| *EPHA10* | *EPHA2* | *EPHA3* | *EPHA5* | *EPHA7* | *EPHB1* | *EPHB6* |
| *EPO* | *EPOR* | *EPS15* | *ERBB2* | *ERBB3* | *ERBB4* | *ERC1* |
| *ERCC1* | *ERCC2* | *ERCC3* | *ERCC4* | *ERCC5* | *ERCC6* | *ERG* |
| *ERLIN2* | *ESR1* | *ETS1* | *ETS2* | *ETV1* | *ETV4* | *ETV5* |
| *ETV6* | *EWSR1* | *EXOSC6* | *EXT1* | *EXT2* | *EYA1* | *EYA2* |
| *EZH2* | *EZR* | *FAF1* | *FAM127C* | *FAM19A2* | *FAM19A5* | *FAM46C* |
| *FAM64A* | *FANCA* | *FANCB* | *FANCC* | *FANCD2* | *FANCE* | *FANCF* |
| *FANCG* | *FANCI* | *FANCL* | *FANCM* | *FAS* | *FASLG* | *FBN2* |
| *FBXO11* | *FBXO31* | *FBXW7* | *FCGBP* | *FCGR2B* | *FCRL4* | *FEN1* |

| | | | | | | |
|---|---|---|---|---|---|---|
| *FEV* | *FGF1* | *FGF10* | *FGF13* | *FGF14* | *FGF19* | *FGF2* |
| *FGF23* | *FGF3* | *FGF4* | *FGF6* | *FGF8* | *FGF9* | *FGFR1* |
| *FGFR1OP* | *FGFR1OP2* | *FGFR2* | *FGFR3* | *FGFR4* | *FH* | *FHIT* |
| *FHL2* | *FIGF* | *FIP1L1* | *FLCN* | *FLI1* | *FLNA* | *FLNC* |
| *FLT1* | *FLT3* | *FLT3LG* | *FLT4* | *FLYWCH1* | *FNBP1* | *FOS* |
| *FOSB* | *FOSL1* | *FOXL2* | *FOXO1* | *FOXO3* | *FOXO4* | *FOXP1* |
| *FRK* | *FRMPD4* | *FRS2* | *FRYL* | *FSTL3* | *FUS* | *FUT1* |
| *FZD10* | *FZD2* | *FZD3* | *FZD6* | *FZD7* | *FZD8* | *GAB1* |
| *GABRG2* | *GADD45B* | *GANAB* | *GAS1* | *GAS5* | *GAS7* | *GATA1* |
| *GATA2* | *GATA3* | *GATA6* | *GBP2* | *GDF6* | *GFAP* | *GHR* |
| *GID4* | *GIT2* | *GLI1* | *GLI3* | *GMPS* | *GNA11* | *GNA12* |
| *GNA13* | *GNAI1* | *GNAQ* | *GNAS* | *GNG4* | *GOLGA5* | *GOPC* |
| *GOSR1* | *GOT1* | *GPC3* | *GPHN* | *GPR124* | *GPR128* | *GPR34* |
| *GRB10* | *GRB2* | *GRHPR* | *GRID1* | *GRIN2A* | *GRIN2B* | *GRM1* |
| *GRM3* | *GSK3B* | *GSN* | *GSTT1* | *GTF2I* | *GTSE1* | *H2AFX* |
| *H3F3A* | *HAS2* | *HDAC1* | *HDAC2* | *HDAC3* | *HDAC4* | *HDAC5* |
| *HDAC6* | *HDAC7* | *HECW1* | *HEPH* | *HERPUD1* | *HES1* | *HES5* |

| | | | | | | |
|---|---|---|---|---|---|---|
| *HEY1* | *HGF* | *HHEX* | *HIF1A* | *HIP1* | *HIPK1* | *HIPK2* |
| *HIST1H1C* | *HIST1H1D* | *HIST1H1E* | *HIST1H2AC* | *HIST1H2AG* | *HIST1H2AL* | *HIST1H2AM* |
| *HIST1H2BC* | *HIST1H2BJ* | *HIST1H2BK* | *HIST1H2BO* | *HIST1H3B* | *HIST1H4I* | *HLF* |
| *HMGA1* | *HMGA2* | *HMGB1* | *HMGN2P46* | *HNF1A* | *HNRNPA2B1* | *HOOK3* |
| *HOXA10* | *HOXA11* | *HOXA13* | *HOXA3* | *HOXA9* | *HOXC11* | *HOXC13* |
| *HOXD11* | *HOXD13* | *HOXD9* | *HRAS* | *HSP90AA1* | *HSP90AB1* | *HSPA1A* |
| *HSPA2* | *HSPA4* | *HSPA5* | *HTRA1* | *HUWE1* | *IBSP* | *ICAM1* |
| *ICK* | *ID1* | *ID3* | *ID4* | *IDH1* | *IDH2* | *IFNG* |
| *IFRD1* | *IGF1* | *IGF1R* | *IGFBP2* | *IGFBP3* | *IKBKB* | *IKBKE* |
| *IKZF1* | *IKZF2* | *IKZF3* | *IL12RB2* | *IL13* | *IL13RA2* | *IL15* |
| *IL1B* | *IL1R1* | *IL1RAP* | *IL2* | *IL21R* | *IL2RA* | *IL3* |
| *IL6* | *IL7R* | *INHBA* | *INPP4A* | *INPP4B* | *INPP5A* | *INPP5D* |
| *IQCG* | *IRF1* | *IRF2BP2* | *IRF4* | *IRF8* | *IRS1* | *IRS2* |
| *IRS4* | *ITGA5* | *ITGA7* | *ITGA8* | *ITGAV* | *ITGB3* | *ITK* |
| *ITPKA* | *JAG2* | *JAK1* | *JAK2* | *JAK3* | *JARID2* | *JAZF1* |
| *JUN* | *KALRN* | *KANK1* | *KAT2B* | *KAT6A* | *KAT6B* | *KCNB1* |
| *KDM1A* | *KDM2B* | *KDM4C* | *KDM5A* | *KDM5C* | *KDM6A* | *KDR* |

| | | | | | | |
|---|---|---|---|---|---|---|
| *KDSR* | *KEAP1* | *KIAA0232* | *KIAA1524* | *KIAA1549* | *KIAA1598* | *KIF5B* |
| *KIT* | *KLF4* | *KLHL6* | *KLK2* | *KLK7* | *KMT2A* | *KMT2B* |
| *KMT2C* | *KMT2D* | *KPNB1* | *KRAS* | *KSR1* | *KTN1* | *LAMA1* |
| *LAMA5* | *LAMP2* | *LASP1* | *LCK* | *LCP1* | *LEF1* | *LEFTY2* |
| *LFNG* | *LGALS3* | *LGR5* | *LHFP* | *LHX2* | *LHX4* | *LIFR* |
| *LINC00598* | *LINC00982* | *LINGO2* | *LMBRD1* | *LMO1* | *LMO2* | *LMO7* |
| *LNP1* | *LOX* | *LPAR1* | *LPP* | *LPXN* | *LRIG3* | *LRMP* |
| *LRP1B* | *LRP5* | *LRPPRC* | *LRRC37B* | *LRRC59* | *LRRC7* | *LRRK2* |
| *LTBP1* | *LYL1* | *LYN* | *MACROD1* | *MAD2L1* | *MADD* | *MAF* |
| *MAFB* | *MAGED1* | *MAGEE1* | *MALAT1* | *MALT1* | *MAML1* | *MAML2* |
| *MAP2* | *MAP2K1* | *MAP2K2* | *MAP2K3* | *MAP2K4* | *MAP2K5* | *MAP2K6* |
| *MAP2K7* | *MAP3K1* | *MAP3K14* | *MAP3K6* | *MAP3K7* | *MAPK1* | *MAPK3* |
| *MAPK8* | *MAPK8IP2* | *MAPK9* | *MAPRE1* | *MATK* | *MAX* | *MB21D2* |
| *MBNL1* | *MBTD1* | *MCL1* | *MDC1* | *MDH1* | *MDM2* | *MDM4* |
| *MDS2* | *MEAF6* | *MECOM* | *MED12* | *MEF2B* | *MEF2C* | *MEF2D* |
| *MELK* | *MEN1* | *MET* | *METTL18* | *METTL7B* | *MFNG* | *MGEA5* |
| *MGMT* | *MIB1* | *MIPOL1* | *MITF* | *MKI67* | *MKL1* | *MKL2* |

| | | | | | | |
|---|---|---|---|---|---|---|
| *MLF1* | *MLH1* | *MLLT1* | *MLLT10* | *MLLT11* | *MLLT3* | *MLLT4* |
| *MLLT6* | *MMP7* | *MMP9* | *MN1* | *MNAT1* | *MNX1* | *MPL* |
| *MRE11A* | *MSH2* | *MSH3* | *MSH6* | *MSI2* | *MSN* | *MTCP1* |
| *MTOR* | *MTUS2* | *MUC1* | *MUTYH* | *MYB* | *MYBL1* | *MYC* |
| *MYCL* | *MYCN* | *MYD88* | *MYH11* | *MYH9* | *MYO18A* | *MYO1F* |
| *NAB2* | *NACA* | *NAPA* | *NAV3* | *NBEAP1* | *NBN* | *NBR1* |
| *NCAM1* | *NCKIPSD* | *NCOA1* | *NCOA2* | *NCOA3* | *NCOA4* | *NCOR2* |
| *NCSTN* | *NDC80* | *NDE1* | *NDRG1* | *NDUFAF1* | *NEDD4* | *NEURL1* |
| *NF1* | *NF2* | *NFATC1* | *NFATC2* | *NFE2L2* | *NFIB* | *NFKB1* |
| *NFKB2* | *NFKBIA* | *NGF* | *NGFR* | *NIN* | *NIPBL* | *NKX2-1* |
| *NKX2-5* | *NOD1* | *NODAL* | *NONO* | *NOS3* | *NOTCH1* | *NOTCH2* |
| *NOTCH3* | *NOTCH4* | *NPM1* | *NPM2* | *NR3C1* | *NR4A3* | *NR6A1* |
| *NRAS* | *NSD1* | *NT5C2* | *NTF3* | *NTF4* | *NTRK1* | *NTRK2* |
| *NTRK3* | *NUMA1* | *NUP107* | *NUP214* | *NUP93* | *NUP98* | *NUTM1* |
| *NUTM2A* | *NUTM2B* | *OFD1* | *OLIG1* | *OLIG2* | *OLR1* | *OMD* |
| *P2RY8* | *PAFAH1B2* | *PAG1* | *PAK1* | *PAK3* | *PAK6* | *PAK7* |
| *PALB2* | *PAPPA* | *PASK* | *PATZ1* | *PAX3* | *PAX5* | *PAX7* |

| | | | | | | |
|---|---|---|---|---|---|---|
| *PAX8* | *PBRM1* | *PBX1* | *PC* | *PCBP1* | *PCLO* | *PCM1* |
| *PCNA* | *PCSK7* | *PDCD1* | *PDCD11* | *PDCD1LG2* | *PDE4DIP* | *PDGFA* |
| *PDGFB* | *PDGFD* | *PDGFRA* | *PDGFRB* | *PDK1* | *PEG3* | *PER1* |
| *PFDN5* | *PHB* | *PHF1* | *PHF23* | *PHF6* | *PHOX2B* | *PI4KA* |
| *PICALM* | *PIK3CA* | *PIK3CB* | *PIK3CD* | *PIK3CG* | *PIK3R1* | *PIK3R2* |
| *PIM1* | *PKM* | *PLA2G2A* | *PLA2G5* | *PLAG1* | *PLAT* | *PLAU* |
| *PLCB1* | *PLCB4* | *PLCG1* | *PLCG2* | *PLEKHM2* | *PML* | *PMS1* |
| *PMS2* | *POFUT1* | *POLD1* | *POLD4* | *POLR2H* | *POM121* | *POMGNT1* |
| *POSTN* | *POT1* | *POU2AF1* | *POU5F1* | *PPAP2B* | *PPARG* | *PPARGC1A* |
| *PPFIA2* | *PPFIBP1* | *PPM1D* | *PPP1CB* | *PPP1R13B* | *PPP1R13L* | *PPP2CB* |
| *PPP2R1A* | *PPP2R1B* | *PPP2R2B* | *PPP2R4* | *PPP3CA* | *PPP3CB* | *PPP3CC* |
| *PPP3R1* | *PPP3R2* | *PPP4C* | *PQLC3* | *PRCC* | *PRDM1* | *PRDM16* |
| *PRDM7* | *PRF1* | *PRG2* | *PRICKLE1* | *PRKACA* | *PRKACG* | *PRKAR1A* |
| *PRKCA* | *PRKCB* | *PRKCD* | *PRKCG* | *PRKDC* | *PRKG2* | *PRMT1* |
| *PRMT8* | *PROM1* | *PRRX1* | *PRRX2* | *PRSS8* | *PSD3* | *PSEN1* |
| *PSIP1* | *PSMD2* | *PTBP1* | *PTCH1* | *PTCRA* | *PTEN* | *PTGS2* |
| *PTK2* | *PTK2B* | *PTK7* | *PTPN11* | *PTPN2* | *PTPN6* | *PTPRA* |

| | | | | | | |
|---|---|---|---|---|---|---|
| *PTPRK* | *PTPRO* | *PTPRR* | *PTTG1* | *PVT1* | *RABEP1* | *RAC1* |
| *RAC2* | *RAC3* | *RAD21* | *RAD50* | *RAD51* | *RAD51B* | *RAD51C* |
| *RAD51D* | *RAD52* | *RAF1* | *RALGDS* | *RANBP17* | *RANBP2* | *RAP1GDS1* |
| *RARA* | *RASAL1* | *RASGEF1A* | *RASGRF1* | *RASGRF2* | *RASGRP1* | *RB1* |
| *RBM15* | *RBM6* | *RCHY1* | *RCOR1* | *RCSD1* | *RECQL4* | *REEP3* |
| *RELA* | *RELN* | *RERG* | *RET* | *RGS7* | *RHBDF2* | *RHOA* |
| *RHOD* | *RHOH* | *RICTOR* | *RLTPR* | *RMI2* | *RNF213* | *RNF43* |
| *ROBO1* | *ROBO2* | *ROS1* | *RPA3* | *RPL22* | *RPN1* | *RPN2* |
| *RPS21* | *RPS6KA1* | *RPS6KA2* | *RPS6KA3* | *RPTOR* | *RREB1* | *RRM1* |
| *RRM2B* | *RTEL1* | *RTN3* | *RUNX1* | *RUNX1T1* | *RUNX2* | *RYR3* |
| *S1PR2* | *SARNP* | *SBDS* | *SCN8A* | *SDC4* | *SDHA* | *SDHAF2* |
| *SDHB* | *SDHC* | *SDHD* | *SEC31A* | *SEPT2* | *SEPT5* | *SEPT6* |
| *SEPT9* | *SERP2* | *SERPINE1* | *SERPINF1* | *SET* | *SETBP1* | *SETD2* |
| *SETD7* | *SF3B1* | *SFPQ* | *SFRP2* | *SFRP4* | *SGK1* | *SGPP2* |
| *SH2D5* | *SH3BP1* | *SH3D19* | *SH3GL1* | *SH3GL2* | *SHC1* | *SHC2* |
| *SIK3* | *SIN3A* | *SIRT1* | *SKP2* | *SLC1A2* | *SLC34A2* | *SLC45A3* |
| *SLC7A5* | *SLCO1B3* | *SLX4* | *SMAD2* | *SMAD3* | *SMAD4* | *SMAD6* |

| | | | | | | |
|---|---|---|---|---|---|---|
| *SMAP1* | *SMARCA1* | *SMARCA4* | *SMARCA5* | *SMARCB1* | *SMC1A* | *SMC3* |
| *SMO* | *SNAPC3* | *SNCG* | *SNHG5* | *SNW1* | *SNX29* | *SNX9* |
| *SOCS1* | *SOCS2* | *SOCS3* | *SOD2* | *SORBS2* | *SORT1* | *SOS1* |
| *SOX10* | *SOX11* | *SOX2* | *SP1* | *SP3* | *SPECC1* | *SPEN* |
| *SPOP* | *SPP1* | *SPRY2* | *SPRY4* | *SPTAN1* | *SPTBN1* | *SQSTM1* |
| *SRC* | *SRF* | *SRGAP3* | *SRRM3* | *SRSF2* | *SRSF3* | *SS18* |
| *SS18L1* | *SSBP2* | *SSX1* | *SSX2* | *SSX4* | *ST6GAL1* | *STAG2* |
| *STAT1* | *STAT3* | *STAT4* | *STAT5A* | *STAT5B* | *STAT6* | *STIL* |
| *STK11* | *STL* | *STRN* | *STX5* | *STYK1* | *SUFU* | *SUGP2* |
| *SULF1* | *SUV39H2* | *SUZ12* | *SYK* | *SYP* | *TACC1* | *TACC2* |
| *TACC3* | *TAF1* | *TAF15* | *TAL1* | *TAL2* | *TAOK1* | *TBL1XR1* |
| *TBX15* | *TCEA1* | *TCF12* | *TCF3* | *TCF7L2* | *TCL1A* | *TCL6* |
| *TCTA* | *TEAD1* | *TEAD2* | *TEAD3* | *TEAD4* | *TEC* | *TENM1* |
| *TERF1* | *TERF2* | *TERT* | *TET1* | *TET2* | *TFAP2A* | *TFDP1* |
| *TFE3* | *TFEB* | *TFG* | *TFPT* | *TFRC* | *TGFB2* | *TGFB3* |
| *TGFBI* | *TGFBR2* | *TGFBR3* | *THADA* | *THBS1* | *THRAP3* | *TIAM1* |
| *TIRAP* | *TLL2* | *TLR4* | *TLX1* | *TLX3* | *TMEM127* | *TMEM230* |

| | | | | | | |
|---|---|---|---|---|---|---|
| TMEM30A | TMPRSS2 | TNC | TNF | TNFAIP3 | TNFRSF10B | TNFRSF10D |
| TNFRSF11A | TNFRSF14 | TNFRSF17 | TNFRSF6B | TOP1 | TOP2A | TOP2B |
| TP53 | TP53BP1 | TP63 | TP73 | TPD52L2 | TPM3 | TPM4 |
| TPO | TPR | TRAF2 | TRAF3 | TRAF5 | TRHDE | TRIM24 |
| TRIM27 | TRIM33 | TRIP11 | TRPS1 | TSC1 | TSC2 | TSHR |
| TTK | TTL | TUSC3 | TYK2 | TYMS | U2AF1 | U2AF2 |
| UBE2B | UBE2C | UFC1 | UFM1 | USP16 | USP42 | USP5 |
| USP6 | USP7 | VCAM1 | VEGFA | VEGFC | VGLL3 | VHL |
| VTI1A | WASF2 | WDFY3 | WDR1 | WDR18 | WDR70 | WDR90 |
| WEE1 | WHSC1 | WHSC1L1 | WIF1 | WISP3 | WNT10A | WNT10B |
| WNT11 | WNT16 | WNT2B | WNT3 | WNT4 | WNT5B | WNT6 |
| WNT7B | WNT8B | WRN | WSB1 | WT1 | WWOX | WWTR1 |
| XBP1 | XIAP | XKR3 | XPA | XPC | XPO1 | XRCC6 |
| YAP1 | YPEL5 | YTHDF2 | YWHAE | YY1AP1 | ZBTB16 | ZC3H7A |
| ZC3H7B | ZFP64 | ZFPM2 | ZFYVE19 | ZIC2 | ZMIZ1 | ZMYM2 |
| ZMYM3 | ZMYND11 | ZNF207 | ZNF217 | ZNF24 | ZNF331 | ZNF384 |
| ZNF444 | ZNF521 | ZNF585B | ZNF687 | ZNF703 | ZRSR2 | |

**C DNA Sequencing**

The NGS procedure was carried out using the SureSelect XT HS2 DNA Reagent Kit (Agilent Technologies, Santa Clara, California, United States) and the SureSelect XT HS and XT Low Input Enzymatic Fragmentation Kit (Agilent Technologies, Santa Clara, California, United States) following the manufacturer's protocol SureSelect XT HS2 DNA System DNA Library Preparation and Target Enrichment for the Illumina Platforms (Agilent Technologies, Santa Clara, California, United States) with a dedicated panel created covering 770 genes leading to oncological diseases (Table..). The prepared libraries were sequenced on the NextSeq 550 platform (Illumina, San Diego, CA) according to the manufacturer's protocol. The process was carried out in paired-end mode covering 300 base pairs using the NextSeq 500/550 Mid Output Kit v2.5 (Illumina, San Diego, CA). Reads were mapped to the GRCh37/hg19 reference gene sequence using the Burrows-Wheeler Aligner (BWA) algorithm. Variant identification was performed using the GATK Variant Calling algorithm. The data were analyzed using Variant Studio v. 3.0 (Illumina San Diego California United States) and Integrative Genomic Viewer.

The list of genes included in targeted DNA Sequencing:

| | | | | | | |
|---|---|---|---|---|---|---|
| *A2ML1* | *CEP152* | *FAN1* | *KMT2C* | *NYNRIN* | *RB1* | *SMC3* |
| *ABCB11* | *CEP164* | *FANCA* | *KMT2D* | *OFD1* | *RB1CC1* | *SMO* |
| *ABCB4* | *CEP57* | *FANCB* | *KRAS* | *OGG1* | *RBBP8* | *SMUG1* |
| *ABL1* | *CEP63* | *FANCC* | *LATS1* | *OPCML* | *RBM8A* | *SOCS2* |
| *ABL2* | *CETN2* | *FANCD2* | *LEF1* | *ORC1* | *RBSN* | *SOS1* |
| *ABRAXAS1* | *CHAF1A* | *FANCE* | *LIG1* | *P2RY12* | *RECK* | *SOS2* |
| *ACD* | *CHEK1* | *FANCF* | *LIG3* | *PALB2* | *RECQL* | *SPRED1* |
| *ADA* | *CHEK2* | *FANCG* | *LIG4* | *PARN* | *RECQL4* | *SPRTN* |
| *ADA2* | *CHIC2* | *FANCI* | *LMO1* | *PARP1* | *RECQL5* | *SRC* |
| *ADAMTS13* | *CIC* | *FANCL* | *LMO2* | *PARP2* | *REL* | *SRGAP1* |
| *AIP* | *CLCN5* | *FANCL* | *LPP* | *PARP3* | *RELA* | *SRGAP2* |
| *AK1* | *CLK2* | *FANCM* | *LRBA* | *PAX3* | *RELN* | *SRP54* |
| *AKT1* | *CLPB* | *FAS* | *LRP2* | *PAX5* | *REST* | *SRP72* |
| *AKT3* | *CLRN1* | *FASLG* | *LRP5* | *PAX6* | *RET* | *SRSF2* |
| *ALDH7A1* | *CMM* | *FBXW7* | *LUC7L2* | *PAX7* | *REV1* | *SRY* |
| *ALK* | *CNOT3* | *FCGR2A* | *LYST* | *PCNA* | *REV3L* | *SS18* |
| *ALKBH1* | *COL7A1* | *FCGR3B* | *LZTR1* | *PCNT* | *RFWD3* | *SSBP2* |
| *ALKBH2* | *CRB2* | *FEN1* | *MAD1L1* | *PDGFB* | *RHAG* | *STAG2* |
| *ALKBH3* | *CREBBP* | *FERMT1* | *MAD2L2* | *PDGFRA* | *RHBDF2* | *STAM* |

| | | | | | | |
|---|---|---|---|---|---|---|
| *ALOX12B* | *CRIPAK* | *FERMT3* | *MAGT1* | *PDGFRB* | *RIF1* | *STAT1* |
| *AMELX* | *CRLF2* | *FGD3* | *MAML2* | *PDGFRL* | *RIT1* | *STAT2* |
| *AMELY* | *CSF1R* | *FGFR1* | *MAP2K1* | *PDPK1* | *RMI1* | *STAT3* |
| *ANK1* | *CSF3R* | *FGFR2* | *MAP2K2* | *PDS5b* | *RMI2* | *STAT4* |
| *ANKRD26* | *CTC1* | *FGFR3* | *MAX* | *PEAR1* | *RMRP* | *STAT5A* |
| *AP3B1* | *CTCF* | *FGTF2H4* | *MBD4* | *PF4* | *RNF168* | *STAT5B* |
| *APC* | *CTLA4* | *FH* | *MCC* | *PF4V1* | *RNF213* | *STAT6* |
| *APEX1* | *CTNNA1* | *FHL1* | *MDC1* | *PHB* | *RNF4* | *STK11* |
| *APEX2* | *CTNNB1* | *FLCN* | *MED12* | *PHF6* | *RNF6* | *STX11* |
| *APLF* | *CTR9* | *FLNA* | *MED12L* | *PHOX2A* | *RNF8* | *STXBP2* |
| *APTX* | *CUX1* | *FLT3* | *MEF2D* | *PHOX2B* | *ROCK2* | *SUFU* |
| *AR* | *CXCR2* | *FOSB* | *MEN1* | *PIAS1* | *ROR2* | *SUZ12* |
| *ARHGAP26* | *CXCR4* | *FOXP1* | *MET* | *PIAS2* | *ROS1* | *TAL1* |
| *ARHGEF12* | *CYCS* | *FOXP3* | *MGMT* | *PIAS3* | *RPA1* | *TBXA2R* |
| *ARID1A* | *CYLD* | *FPR1* | *MINPP1* | *PIAS4* | *RPA2* | *TCF3* |
| *ARID2* | *DCC* | *G6PC3* | *MITF* | *PICALM* | *RPA3* | *TCF7L2* |
| *ASXL1* | *DCLRE1A* | *G6PD* | *MLH1* | *PIK3CA* | *RPA4* | *TCIRG1* |
| *ATM* | *DCLRE1B* | *GALNT12* | *MLH3* | *PIK3CD* | *RPL10* | *TDG* |
| *ATP11C* | *DCLRE1C* | *GATA1* | *MLLT10* | *PIK3CG* | *RPL11* | *TERC* |
| *ATP7B* | *DDB1* | *GATA2* | *MLLT10* | *PIK3R1* | *RPL15* | *TERT* |
| *ATR* | *DDB2* | *GATA3* | *MLLT3* | *PIM1* | *RPL18* | *TET2* |
| *ATRIP* | *DDHD2* | *GBA* | *MMS19* | *PKHD1* | *RPL19* | *TFE3* |
| *ATRX* | *DDR2* | *GCLC* | *MN1* | *PLA2G2A* | *RPL23* | *TFRC* |
| *AURKA* | *DDX3X* | *GDNF* | *MNX1* | *PLAG1* | *RPL26* | *TGFBR1* |
| *AUTS2* | *DDX41* | *GFI1* | *MPL* | *PMS1* | *RPL27* | *TGFBR2* |
| *AXIN1* | *DIAPH1* | *GINS1* | *MPLKIP* | *PMS2* | *RPL31* | *TGFBR3* |
| *AXIN2* | *DIAPH2* | *GJB2* | *MPO* | *PNKP* | *RPL35* | *THPO* |
| *BAP1* | *DIAPH3* | *GNA11* | *MRE11* | *POLB* | *RPL35A* | *TINF2* |
| *BARD1* | *DICER1* | *GNAQ* | *MRE11A* | *POLD1* | *RPL36* | *TJP2* |
| *BAX* | *DIRC3* | *GNB1* | *MRTFA* | *POLE* | *RPL4* | *TLR2* |
| *BCC1* | *DIS3L2* | *GPC3* | *MSH2* | *POLH* | *RPL5* | *TMEM127* |
| *BCL10* | *DKC1* | *GRB2* | *MSH3* | *POLI* | *RPL9* | *TNFRSF14* |

| | | | | | | |
|---|---|---|---|---|---|---|
| *BCL10* | *DLC1* | *GREM1* | *MSH4* | *POLK* | *RPS10* | *TNFRSF6* |
| *BCL11A* | *DLST* | *GTF2H1* | *MSH5* | *POLL* | *RPS15* | *TOPBP1* |
| *BCL11B* | *DMC1* | *GTF2H2* | *MSH6* | *POLM* | *RPS15A* | *TP53* |
| *BCL2* | *DNAJC21* | *GTF2H3* | *MTAP* | *POLN* | *RPS17* | *TP63* |
| *BCL3* | *DNM2* | *GTF2H4* | *MTNR1B* | *POLQ* | *RPS19* | *TPI1* |
| *BCL6* | *DNMT3A* | *GTF2H5* | *MTOR* | *POT1* | *RPS20* | *TPMT* |
| *BCL7A* | *DOCK8* | *H2AX* | *MUC2* | *POU6F2* | *RPS24* | *TREX1* |
| *BCL9* | *DROSHA* | *H3F3A* | *MUS81* | *PPBP* | *RPS26* | *TREX2* |
| *BCOR* | *DTNBP1* | *HAVCR2* | *MUTYH* | *PPM1D* | *RPS27* | *TRIM28* |
| *BCORL1* | *DUSP22* | *HAX1* | *MVK* | *PRF1* | *RPS27A* | *TRIM37* |
| *BCR* | *DUT* | *HBA1* | *MXI1* | *PRKAR1A* | *RPS28* | *TRIP13* |
| *BLM* | *EBF1* | *HBA2* | *MYB* | *PRKCB* | *RPS29* | *TRPM7* |
| *BLOC1S3* | *ECT2L* | *HBB* | *MYBL1* | *PRKCD* | *RPS7* | *TSC1* |
| *BMP4* | *EED* | *HEATR3* | *MYC* | *PRKDC* | *RPSA* | *TSC2* |
| *BMPR1A* | *EFL1* | *HELQ* | *MYCBP2* | *PRKN* | *RRAS* | *TSR2* |
| *BPGM* | *EGFR* | *HFE* | *MYCN* | *PROM1* | *RSPO1* | *TUBB1* |
| *BRAF* | *EGLN1* | *HGD* | *MYD88* | *PRPF19* | *RTEL1* | *TYK2* |
| *BRCA1* | *EIF2AK3* | *HLTF* | *MYH9* | *PRPF8* | *RUNX1* | *U2AF1* |
| *BRCA2* | *ELANE* | *HMBS* | *MYLK2* | *PRSS1* | *RYR2* | *U2AF2* |
| *BRIP1* | *EME1* | *HMCN1* | *NABP2* | *PTCH1* | *SAMD9* | *UBE2T* |
| *BTG1* | *EME2* | *HMGA2* | *NAF1* | *PTCH2* | *SAMD9L* | *UBE2V2* |
| *BTK* | *EP300* | *HMMR* | *NBEAL2* | *PTEN* | *SBDS* | *UNC13D* |
| *BUB1* | *EPAS1* | *HNF1A* | *NBN* | *PTGS1* | *SDHA* | *UROD* |
| *BUB1B* | *EPB41* | *HOXB13* | *NCOA2* | *PTPN11* | *SDHAF2* | *USH2A* |
| *BUB1B* | *EPB42* | *HPS1* | *NCOR1* | *PTPN12* | *SDHB* | *USP7* |
| *BUB3* | *EPCAM* | *HPS4* | *NCOR2* | *PTPN2* | *SDHC* | *UVSSA* |
| *C9* | *EPHA2* | *HRAS* | *NF1* | *PTPN6* | *SDHD* | *VHL* |
| *CALR* | *EPHB2* | *HUS1* | *NF2* | *PTPRC* | *SERPINA1* | *VPS13B* |
| *CASP10* | *EPO* | *HYOU1* | *NFIX* | *PTPRD* | *SETBP1* | *VPS45* |
| *CASP8* | *EPOR* | *IDH1* | *NHEJ1* | *PTPRJ* | *SETD1B* | *VWF* |
| *CASP8AP2* | *ERBB2* | *IDH2* | *NHP2* | *PTPRT* | *SETD2* | *WAS* |
| *CBFB* | *ERBB3* | *IGF2R* | *NIPBL* | *RAB27A* | *SETMAR* | *WRN* |

| | | | | | | |
|---|---|---|---|---|---|---|
| *CBL* | *ERBB4* | *IKZF1* | *NOP10* | *RABGGTA* | *SF1* | *WT1* |
| *CCND1* | *ERCC1* | *IKZF3* | *NOS3* | *RAC1* | *SF3A1* | *WWOX* |
| *CCND2* | *ERCC2* | *IL7R* | *NOTCH1* | *RAC2* | *SF3B1* | *XAB2* |
| *CCND3* | *ERCC3* | *IRF1* | *NOTCH2* | *RAD17* | *SGK1* | *XIAP* |
| *CCNH* | *ERCC4* | *ITK* | *NOTCH3* | *RAD18* | *SH2B1* | *XPA* |
| *CD27* | *ERCC5* | *JAGN1* | *NOTCH4* | *RAD21* | *SH2B3* | *XPC* |
| *CD274(PD-L1)* | *ERCC6* | *JAK1* | *NPHP3* | *RAD23A* | *SH2D1A* | *XRCC1* |
| *CD36* | *ERCC8* | *JAK2* | *NPM1* | *RAD23B* | *SH3GL1* | *XRCC2* |
| *CD40LG* | *ERG* | *JAK3* | *NQO2* | *RAD50* | *SHC1* | *XRCC2* |
| *CD79B* | *ESR1* | *JMJD1C* | *NR4A3* | *RAD51* | *SHOC2* | *XRCC3* |
| *CD96* | *ETNK1* | *KAT6A* | *NRAS* | *RAD51A* | *SHPRH* | *XRCC4* |
| *CDC73* | *ETS1* | *KCNQ1OT1* | *NRG3* | *RAD51B* | *SLC22A18* | *XRCC5* |
| *CDC73* | *ETV6* | *KDM3B* | *NSD1* | *RAD51C* | *SLC25A13* | *XRCC6* |
| *CDH1* | *EWSR1* | *KDM3B* | *NSD2* | *RAD51D* | *SLX1B* | *ZBTB33* |
| *CDHR1* | *EXO1* | *KDM6A* | *NT5C2* | *RAD51D* | *SLX4* | *ZBTB7A* |
| *CDK4* | *EXT1* | *KDM6B* | *NTHL1* | *RAD52* | *SMAD4* | *ZFHX3* |
| *CDK7* | *EXT2* | *KDR* | *NTRK1* | *RAD54B* | *SMAD7* | *ZNF384* |
| *CDKN1B* | *EYS* | *KIF1B* | *NTRK2* | *RAD54L* | *SMARCA2* | *ZNF91* |
| *CDKN1C* | *EZH2* | *KIF1Bβ* | *NTRK3* | *RAD9A* | *SMARCA4* | *ZRSR2* |
| *CDKN2A* | *F8* | *KIT* | *NUDT1* | *RAF1* | *SMARCAL1* | |
| *CDKN2B* | *F9* | *KLF6* | *NUP214* | *RAG1* | *SMARCB1* | |
| *CEBPA* | *FAAP20* | *KLHDC8B* | *NUP98* | *RAG2* | *SMARCD2* | |
| *CENPJ* | *FAAP24* | *KMT2A* | *NUTM1* | *RANBP17* | *SMARCE1* | |
| *CENPP* | *FAH* | *KMT2B* | *NUTM2B-AS1* | *RARA* | *SMC1A* | |

**D Direct sequencing**

Causative and possibly candidate variants detected through NGS were verified by direct Sanger sequencing in the proband. Primers were designed based on sequences available at https://genome.ucsc.edu/. Each primer pair was approximately 20 nucleotides in length. Primer quality was evaluated using NetPrimer (https://www.premierbiosoft.com/netprimer/). We selected primers with a rating above 90%, an optimal melting temperature (Tm) around 60°C, and a temperature difference between the forward and reverse primers not exceeding 3°C, ideally within 1°C. Additional selection criteria included ΔG values no lower than –6 kcal/mol for cross-dimers, –5 kcal/mol for self-dimers, and –1 kcal/mol for hairpins.

Ordered primers were optimized using a temperature gradient, both with and without Q-solution, using HotStarTaq DNA Polymerase (250 U) (QIAGEN, Cat. no. 203203). PCR products were purified using the Clean-up Concentration Kit (A&A Biotechnology; Poland) according to the manufacturer's protocol. Subsequently, the forward and reverse strands were independently labeled using ddNTPs. After labeling, sequencing reactions were purified with ExTerminator (A&A Biotechnology, Poland). Biosystems), followed by purification with the BigDye XTerminator™ Purification Kit (Applied Biosystems). Capillary electrophoresis was performed using the 3500Dx Genetic Analyzer (Applied Biosystems). Final sequence data were analyzed using Sequencher v5.0 software.

**Supplemental Figure 1.**

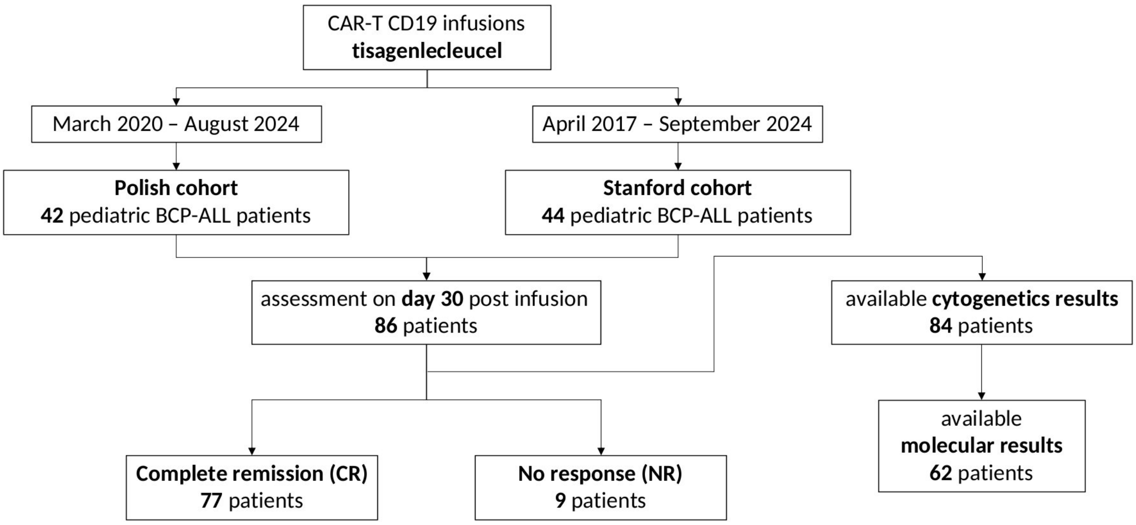


**Supplemental Figure 1.**

**Study selection for univariate analysis of complete response (CR; defined as 30 days post-CAR-T CD19 infusion) and availability of molecular and cytogenetic data.**

**Supplemental Figure 2.**

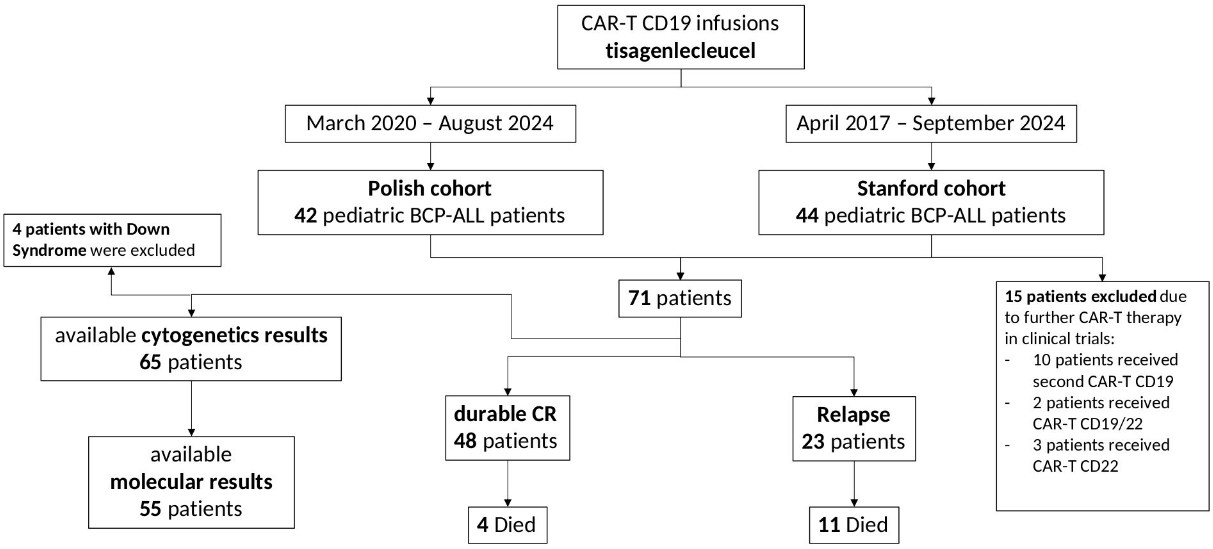


**Supplemental Figure 2.**

**Study selection for overall survival (OS) and leukemia-free survival (LFS) analysis in 72 patients treated with CAR-T CD19 therapy (tisagenlecleucel). Fourteen patients who received additional or different CAR-T constructs (e.g., CD19/22) were excluded. Inclusion flow for OS/LFS analysis and availability of molecular and cytogenetic data.**

**Supplemental Table 1. Univariate analysis of CR rates among subgroups.**

Among 86 patients receiving CAR-T CD19 infusion, those with BM blasts >20% had significantly lower CR rates than those with ≤20% (53.8% vs. 95.9%, p<0.0001). CR rates by patient characteristics were assessed using the Chi-square test.

| Variable | Category | CR | NR | Number of patients (%) | CR Rate (95% CI) | p Value |
|---|---|---|---|---|---|---|
| **Gender** | F | 34 | 4 | 38 (44.2%) | 89.5% (75.9-95.8%) | 1 |
| | M | 43 | 5 | 48 (55.8%) | 89.6% (77.8-95.5%) | |
| **Age [years]** | <10 | 33 | 5 | 38 (44.2%) | 86.8% (72.7-94.2%) | 0,71 |
| | ≥10 | 44 | 4 | 48 (55.8%) | 91.7% (80.4-96.7%) | |
| **BCP-ALL Subtype** | B-I | 2 | 1 | 3 (3.5%) | 66.7% (20.8-93.9%) | 0,55 |
| | B-II | 22 | 2 | 24 (27.9%) | 91.7% (74.2-97.7%) | |
| | B-III | 12 | 1 | 13 (15.1%) | 92.3% (66.7-98.6%) | |
| | MPAL | 1 | 0 | 1 (1.2%) | 100% (20.7-100%) | |
| **Group risk** | HR | 51 | 5 | 56 (65.1%) | 91.1% (80.7-96.1%) | 0,9801 |
| | no HR | 22 | 3 | 25 (29.1%) | 88% (70-95.8%) | |
| **Burden of disease (MRD)** | negative | 41 | 2 | 43 (50%) | 95.3% (84.5-98.7%) | 0,1265 |
| | positive | 33 | 7 | 40 (46.5%) | 82.5% (68.1-91.3%) | |
| **BM blasts by morphology** | <20% | 70 | 3 | 73 (84.9%) | 95.9% (88.6-98.6%) | <0.0001 |
| | >=20% | 7 | 6 | 13 (15.1%) | 53.8% (29.1-76.8%) | |
| **Number of relapses** | refractory | 16 | 2 | 18 (20.9%) | 88.9% (67.2-96.9%) | 0,9728 |
| | 1 | 34 | 4 | 38 (44.2%) | 89.5% (75.9-95.8%) | |
| | 2 | 23 | 3 | 26 (30.2%) | 88.5% (71-96%) | |
| | 3 | 2 | 0 | 2 (2.3%) | 100% (34.2-100%) | |
| | 4 | 2 | 0 | 2 (2.3%) | 100% (34.2-100%) | |
| **Treatment before CAR-T CD19 infusion** | Blinatumomab | 18 | 3 | 21 (24.4%) | 85.7% (65.4-95%) | 0,8564 |
| | Inotuzumab | 21 | 4 | 25 (29.1%) | 84% (65.3-93.6%) | 0,493 |
| | HSCT | 31 | 1 | 32 (37.2%) | 96.9% (84.3-99.4%) | 0,1778 |
| **CRS [grade]** | 0-2 | 74 | 8 | 82 (95.3%) | 90.2% (81.9-95%) | 0.3627* |
| | 3-4 | 3 | 1 | 4 (4.7%) | 75% (30.1-95.4%) | |
| | Tocilizumab | 21 | 5 | 26 (30.2%) | 80.8% (62.1-91.5%) | 0,1724 |
| **ICANS [grade]** | 0-2 | 74 | 9 | 83 (96.5%) | 89.2% (80.7-94.2%) | 1* |
| | 3-4 | 3 | 0 | 3 (3.5%) | 100% (43.9-100%) | |
| **cytogenetic risk group** | HRG | 24 | 5 | 29 (33.7%) | 82.8% (65.5-92.4%) | 0,3579 |
| | IRG | 34 | 3 | 37 (43%) | 91.9% (78.7-97.2%) | |
| | LRG | 17 | 1 | 18 (20.9%) | 94.4% (74.2-99%) | |
| **cytogenetic subtype** | PAX5r | 3 | 2 | 5 (5.8%) | 60% (23.1-88.2%) | 0.0866* |
| | no PAX5r | 72 | 7 | 79 (91.9%) | 91.1% (82.8-95.6%) | |
| | KMT2Ar | 2 | 2 | 4 (4.7%) | 50% (15-85%) | 0.05511* |
| | no KMT2Ar | 73 | 7 | 80 (93%) | 91.2% (83-95.7%) | |
| | *ETV6::RUNX1* | 7 | 1 | 8 (9.3%) | 87.5% (52.9-97.8%) | 1 |
| | no *ETV6::RUNX1* | 68 | 8 | 76 (88.4%) | 89.5% (80.6-94.6%) | |

| | | | | | | |
|---|---|---|---|---|---|---|
| | Hyperdiploid | 10 | 0 | 10 (11.6%) | 100% (72.2-100%) | 0,5336 |
| | no Hyperdiploid | 65 | 9 | 74 (86%) | 87.8% (78.5-93.5%) | |
| | Ph-like | 8 | 1 | 9 (10.5%) | 88.9% (56.5-98%) | 1 |
| | no Ph-like | 67 | 8 | 75 (87.2%) | 89.3% (80.3-94.5%) | |
| **gene mutations** | IKZF1 plus (+) | 8 | 3 | 11 (12.8%) | 72.7% (43.4-90.3%) | 0,2839 |
| | IKZF1 plus (-) | 46 | 5 | 51 (59.3%) | 90.2% (79-95.7%) | |
| | IKZF1 del (+) | 15 | 1 | 16 (18.6%) | 93.8% (71.7-98.9%) | 0,625 |
| | IKZF1 del (-) | 39 | 7 | 46 (53.5%) | 84.8% (71.8-92.4%) | |
| | CDKN2A (+) | 6 | 1 | 7 (8.1%) | 85.7% (48.7-97.4%) | 1 |
| | CDKN2A (-) | 48 | 7 | 55 (64%) | 87.3% (76-93.7%) | |
| | CDKN2B (+) | 4 | 1 | 5 (5.8%) | 80% (37.6-96.4%) | 1 |
| | CDKN2B (-) | 50 | 7 | 57 (66.3%) | 87.7% (76.8-93.9%) | |
| | CREBBP (+) | 5 | 0 | 5 (5.8%) | 100% (56.6-100%) | 1* |
| | CREBBP (-) | 49 | 8 | 57 (66.3%) | 86% (74.7-92.7%) | |
| | ETV6 (+) | 5 | 0 | 5 (5.8%) | 100% (56.6-100%) | 1* |
| | ETV6 (-) | 49 | 8 | 57 (66.3%) | 86% (74.7-92.7%) | |
| | KRAS (+) | 14 | 4 | 18 (20.9%) | 77.8% (54.8-91%) | 0,3258 |
| | KRAS (-) | 40 | 4 | 44 (51.2%) | 90.9% (78.8-96.4%) | |
| | NRAS (+) | 8 | 2 | 10 (11.6%) | 80% (49-94.3%) | 0,829 |
| | NRAS (-) | 46 | 6 | 52 (60.5%) | 88.5% (77-94.6%) | |
| | TP53 (+) | 4 | 2 | 6 (7%) | 66.7% (30-90.3%) | 0,3524 |
| | TP53 (-) | 50 | 6 | 56 (65.1%) | 89.3% (78.5-95%) | |
| | NRAS/KRAS (+) | 19 | 5 | 24 (27.9%) | 79.2% (59.5-90.8%) | 0,2751 |
| | NRAS/KRAS (-) | 35 | 3 | 38 (44.2%) | 92.1% (79.2-97.3%) | |
| | CDKN2A/B (+) | 7 | 1 | 8 (9.3%) | 87.5% (52.9-97.8%) | 1 |
| | CDKN2A/B (-) | 47 | 7 | 54 (62.8%) | 87% (75.6-93.6%) | |
| | TP53/NRAS/KRAS (+) | 22 | 5 | 27 (31.4%) | 81.5% (63.3-91.8%) | 0,4375 |
| | TP53/NRAS/KRAS (-) | 32 | 3 | 35 (40.7%) | 91.4% (77.6-97%) | |
| | Down Syndrome (+) | 6 | 0 | 6 (7%) | 100% (61-100%) | 0,8449 |
| | Down Syndrome (-) | 69 | 9 | 78 (90.7%) | 88.5% (79.5-93.8%) | |
| | Polish group | 38 | 4 | 42 (48.8%) | 90.5% (77.9-96.2%) | 1 |
| | Stanford group | 39 | 5 | 44 (51.2%) | 88.6% (76-95%) | |

**Supplemental Figure 3.**

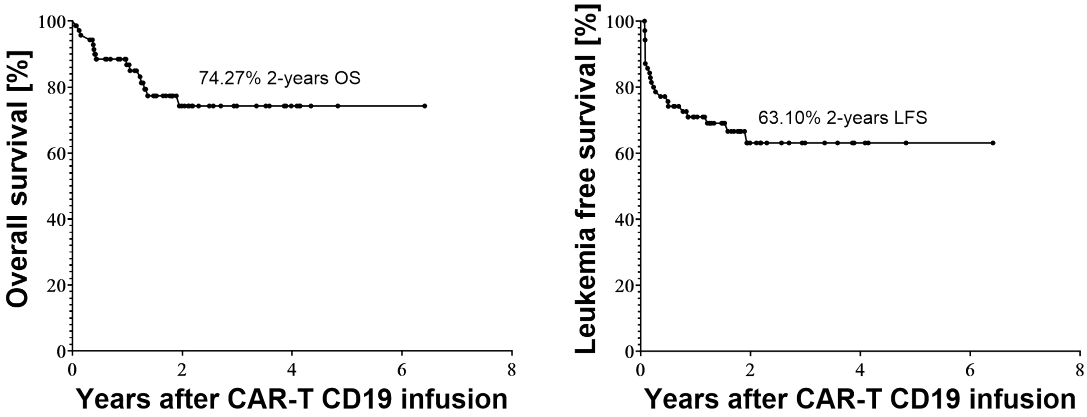


Number at risk

| | | | | |
|---|---|---|---|---|
| OS | 71 | 23 | 6 | 1 |

Number at risk

| | | | | |
|---|---|---|---|---|
| LFS | 71 | 16 | 4 | 1 |

**Supplemental Figure 3.**

2-year OS and LFS were 74.4% and 62.2%, among 71 patients receiving CAR-T CD19 therapy.

**Supplemental Table 2.**

Two-year OS and LFS among subgroups of 71 patients treated with CAR-T CD19 therapy (tisagenlecleucel), after exclusion of 15 patients who received additional or different CAR-T constructs. OS and LFS probabilities were estimated using the Kaplan–Meier method and compared using the Gehan–Breslow–Wilcoxon test. Time-to-event analyses were measured from the date of the first CAR-T cell infusion. Events were censored at disease progression, relapse, death, or last follow-up.

| **Variable** | **Category** | **2-year OS** | **p Value** | **2-year LFS** | **p Value** |
|---|---|---|---|---|---|
| **Gender** | F | 75,40% | 0,7338 | 56,18% | 0,7516 |
| | M | 73,88% | | 66,76% | |
| **Age [years]** | <10 | 67,72% | 0,3626 | 58,43% | 0,965 |
| | ≥10 | 80,62% | | 70,05% | |
| **B-ALL Subtype** | B-I | 66,67% | 0,5595 | 0,00% | 0,5957 |
| | B-II | 87,30% | | 71,59% | |
| | B-III | 76,15% | | 69,23% | |
| **Group risk** | HR | 80,44% | 0,9244 | 61,46% | 0,6145 |
| | no HR | 79,59% | | 70,33% | |
| **Burden of disease (MRD) by NGS/PCR** | negative | 85,93% | 0,0615 | 75,96% | 0,0176 |
| | positive | 68,57% | | 56,25% | |
| **BM blasts by morphology** | <20% | 82,90% | <0.0001 | 70,96% | <0.0001 |
| | >=20% | 30,00% | | 20,00% | |
| **Number of relapses** | refractory | 56,25% | 0,8671 | 34,09% | 0,89 |
| | 1 | 79,44% | | 69,41% | |
| | >=2 | 74,97% | | 62,22% | |
| **Treatment before CAR-T CD19 infusion** | Blinatumomab | 61,91% | 0,2302 | 65,63% | 0,8292 |
| | Inotuzumab | 63,80% | 0,0121 | 45,00% | 0,1139 |
| | HSCT | 82,74% | 0,3044 | 77,82% | 0,0307 |
| **CRS [grade]** | 0-1 | 80,48% | 0,0201 | 67,65% | 0,0481 |
| | 2--4 | 52,38% | | 47,62% | |
| | Tocilizumab | 62,90% | 0,0334 | 61,11% | 0,224 |
| **ICANS [grade]** | 0-2 | 75,00% | 0,1678 | 63,50% | 0,5716 |
| | 3-4 | 50,00% | | 50,00% | |
| **cytogenetic risk group*** | HRG | 70,17% | 0,4138 | 39,57% | 0,1142 |
| | IRG | 80,49% | | 67,29% | |
| | LRG | 85,12% | | 78,57% | |
| **cytogenetic subtype** (vs B-other) | *ETV6::RUNX1* | 85,71% | 0,7585 | 85,71% | 0,4488 |
| | Hyperdiploid | 85,71% | 0,7535 | 71,43% | 0,7542 |
| | Ph-like | 55,56% | 0,1739 | 83,33% | 0,5262 |
| | PAX5r | 50,00% | 0,0289 | 25,00% | 0,0886 |
| | KMT2Ar | 50,00% | 0,0771 | 25,00% | 0,0967 |
| **gene mutations** | IKZF1 plus | 76,19% | 0,8452 | 44,44% | 0,5493 |
| | IKZF1 del | 93,75% | 0,1496 | 75,00% | 0,1101 |

| | | | | | |
|---|---|---|---|---|---|
| | CDKN2A | 50,00% | 0,7099 | 0,00% | 0,7697 |
| | CDKN2B | 0,00% | 0,3862 | 100,00% | 0,5485 |
| | CREBBP | 80,00% | 0,9271 | 80,00% | 0,5543 |
| | ETV6 | 66,67% | 0,6363 | 80,00% | 0,4309 |
| | KRAS | 59,23% | 0,0239 | 56,25% | 0,0833 |
| | NRAS | 77,78% | 0,6222 | 27,78% | 0,2145 |
| | TP53 | 60,00% | 0,1269 | 60,00% | 0,4898 |
| | NRAS/KRAS | 64,14% | 0,0222 | 45,40% | 0,0402 |
| | CDKN2A/B | 50,00% | 0,7099 | 0,00% | 0,7697 |
| | TP53/NRAS/KRAS | 69,03% | 0,0675 | 54,44% | 0,1389 |
| | Down Syndrome | 33,33% | 0,7867 | 33,33% | 0,5898 |
| **Treatment post CAR-T CD19 infusion** | HSCT | 68,61% | 0,6577 | 32,73% | 0,0117 |
| | Polish group | 82,75% | 0,6689 | 67,25% | 0,2008 |
| | Stanford group | 60,99% | | 56,53% | |

* Cytogenetic categories were assigned according to Leahy et al. (Blood 2021)[7]. High-risk cytogenetics included KMT2A (MLL) rearrangements, Philadelphia chromosome (Ph+), Ph-like ALL, hypodiploidy, and TCF3/HLF. Favorable cytogenetics included hyperdiploidy and ETV6/RUNX1. Intermediate-risk cytogenetics included iAMP21, IKZF1 deletion, and TCF3/PBX1.

**Supplemental Table 3.**

Association between bone marrow (BM) blast percentage and burden of disease (MRD), cytokine release syndrome (CRS) severity, treatment before CAR-T CD19 infusion, Tocilizumab, PAX5r and RAS. Patients with ≥20% BM blasts were more likely to develop grade 2–4 CRS compared with those with <20% BM blasts. Statistical significance was determined using a chi-square test.

| | | BM blasts by morphology | | p value |
|---|---|---|---|---|
| | | **<20%** | **>=20%** | |
| **CRS** | **grade 0--1** | 53 | 4 | 0.000552 |
| | **grade 2--4** | 8 | 6 | |
| **Treatment before CAR-T CD19 infusion** | **Inotuzumab (+)** | 19 | 2 | 0.474026 |
| | **Inotuzumab (-)** | 42 | 8 | |
| | **HSCT (+)** | 23 | 3 | 0.639227 |
| | **HSCT (-)** | 38 | 7 | |
| **Tocilizumab** | **+** | 17 | 7 | 0.009038 |
| | **-** | 44 | 3 | |
| **PAX5r** | **+** | 4 | 0 | 1 |
| | **-** | 59 | 10 | |
| **RAS** | **+** | 20 | 4 | 0.515 |
| | **-** | 35 | 3 | |

**Supplemental Table 4.**

Comparison of clinical and biological characteristics between patients with available molecular results and patients without it, including demographic data, disease burden and treatment history. P values indicate statistical significance of differences between groups.

| Variable | Category | available molecular results | % of patients | non available molecular results | % of patients | p Value |
|---|---|---|---|---|---|---|
| **Gender** | F | 30 | 48% | 8 | 33% | 0,3083 |
| | M | 32 | 52% | 16 | 67% | |
| **Age [years]** | <10 | 32 | 52% | 9 | 38% | 0,3499 |
| | ≥10 | 30 | 48% | 15 | 63% | |
| **Group risk** | HR | 37 | 60% | 19 | 79% | 0,1474 |
| | no HR | 25 | 40% | 5 | 21% | |
| **Burden of disease (MRD)** | negative | 31 | 50% | 12 | 50% | 1 |
| | positive | 31 | 50% | 12 | 50% | |
| **BM blasts by morphology** | <20% | 55 | 89% | 22 | 92% | 0,9927 |
| | >=20% | 7 | 11% | 2 | 8% | |
| **Number of relapses** | refractory | 13 | 21% | 5 | 21% | 0,7219 |
| | 1 | 30 | 48% | 12 | 50% | |
| | >=2 | 19 | 31% | 11 | 46% | |
| **Treatment before CAR-T CD19 infusion** | Blinatumomab | 15 | 24% | 6 | 25% | 1 |
| | Inotuzumab | 23 | 37% | 2 | 8% | 0,1778 |
| | HSCT | 22 | 35% | 10 | 42% | 1 |
| **CRS [grade]** | 0-1 | 51 | 82% | 19 | 79% | 0,9828 |
| | 2--4 | 11 | 18% | 5 | 21% | |
| | Tocilizumab | 21 | 34% | 5 | 21% | 0,358 |
| **ICANS [grade]** | 0-2 | 60 | 97% | 23 | 96% | 1 |
| | 3-4 | 2 | 3% | 1 | 4% | |
| **HSCT after CAR-T** | | 17 | 27% | 8 | 33% | 0,7818 |

**Supplemental Table 5.**

Comparison of clinical and biological characteristics between RAS-positive and RAS-negative patient groups, including demographic data, disease burden, treatment history, and genetic features. P values indicate statistical significance of differences between groups.

| Variable | Category | RAS(+) | % of patients | RAS(-) | % of patients | p Value |
|---|---|---|---|---|---|---|
| **Gender** | F | 12 | 50% | 18 | 48% | 1 |
| | M | 12 | 50% | 20 | 52% | |
| **Age [years]** | <10 | 12 | 50% | 19 | 50% | 1 |
| | ≥10 | 12 | 50% | 19 | 50% | |
| **Group risk** | HR | 14 | 60% | 23 | 66% | 0.9233 |
| | no HR | 9 | 40% | 12 | 34% | |
| **Burden of disease (MRD)** | negative | 10 | 42% | 21 | 55% | 0.4341 |
| | positive | 14 | 58% | 17 | 45% | |
| **BM blasts by morphology** | <20% | 20 | 83% | 35 | 92% | 0.515 |
| | >=20% | 4 | 17% | 3 | 8% | |
| **Number of relapses** | refractory | 4 | 16% | 9 | 24% | 0.795 |
| | 1 | 12 | 50% | 18 | 47% | |
| | >=2 | 8 | 34% | 11 | 29% | |
| **Treatment before CAR-T CD19 infusion** | Blinatumomab | 6 | 26% | 9 | 25% | 0.7921 |
| | Inotuzumab | 12 | 50% | 11 | 29% | 0.2125 |
| | HSCT | 7 | 29% | 15 | 42% | 0.4771 |
| **CRS [grade]** | 0-1 | 21 | 88% | 30 | 79% | 0.3905 |
| | 2--4 | 3 | 12% | 8 | 21% | |
| | Tocilizumab | 6 | 25% | 15 | % | 0.2408 |
| **ICANS [grade]** | 0-2 | 24 | 100% | 36 | 95% | 0.6858 |
| | 3-4 | 0 | 0% | 2 | 5% | |
| **cytogenetic risk group** | HRG | 7 | 29% | 15 | 39% | 0.5768 |
| | IRG | 12 | 50% | 14 | 37% | |
| | LRG | 5 | 21% | 9 | 24% | |
| **cytogenetic subtype** | *ETV6::RUNX1* | 2 | 8% | 5 | 13% | 0.5114 |
| | Hyperdiploid | 3 | 12% | 4 | 11% | 0.8696 |
| | Ph-like | 1 | 4% | 6 | 16% | 0.159 |
| | KMT2Ar | 1 | 4% | 2 | 5% | 0.8446 |
| | PAX5r | 4 | 16% | 1 | 3% | 0.0480 |
| **gene mutations** | IKZF1 plus | 6 | 25% | 5 | 13% | 0.2345 |
| | IKZF1 del | 5 | 21% | 11 | 29% | 0.477 |
| | CDKN2A | 5 | 21% | 2 | 5% | 0.1402 |
| | CDKN2B | 3 | 12% | 2 | 5% | 0.308 |
| | CREBBP | 3 | 12% | 2 | 5% | 0.308 |
| | ETV6 | 2 | 8% | 3 | 8% | 1 |
| | TP53 | 3 | 12% | 3 | 8% | 0.5982 |
| **HSCT after CAR-T** | | 7 | 29% | 10 | 26% | 0.8064 |

**Supplemental Figure 4.**

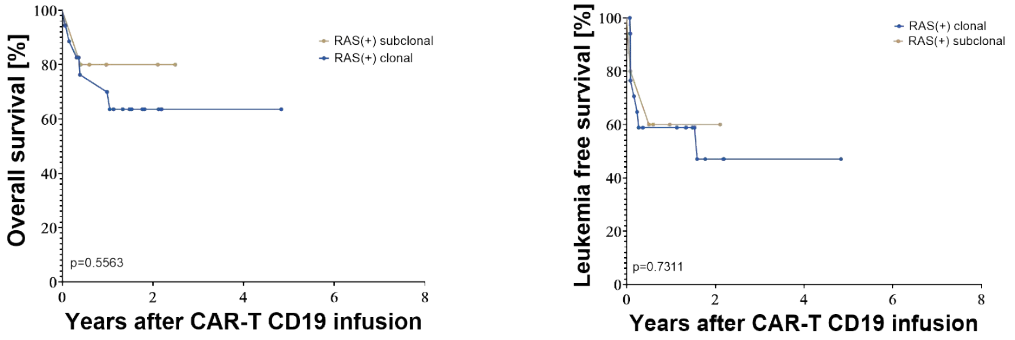


| Number at risk | | | |
|---|---|---|---|
| RAS(+) clonal | 18 | 5 | 1 |
| RAS(+) subclonal | 5 | 2 | 0 |

| Number at risk | | | |
|---|---|---|---|
| RAS(+) clonal | 18 | 4 | 1 |
| RAS(+) subclonal | 5 | 1 | 0 |

Supplemental Figure 4.

OS and LFS in patients with clonal RAS(+) vs subclonal RAS(+) alterations. Clonal RAS mutations were defined as variants with ≥25% variant allele frequency (VAF), whereas subclonal RAS alterations were defined as variants with <25% VAF.
